# Real World SOF/VEL/VOX Retreatment Outcomes and Viral Resistance Analysis for HCV Patients with Prior Failure to DAAs

**DOI:** 10.1101/2020.10.13.20211862

**Authors:** David A Smith, Daniel Bradshaw, Jean Mbisa, Carmen F Manso, David Bibby, Josh Singer, Emma Thomson, Ana Filipe, Elihu Aranday-Cortes, M. Azim Ansari, Anthony Brown, Emma Hudson, Jennifer Benselin, Brendan Healy, Phil Troke, STOP-HCV Consortium, HCV Research UK, John McLauchlan, Eleanor Barnes, William L Irving

**Author notes:** Corresponding Author: Daniel Bradshaw, National Infection Service, Public Health England, Colindale, 61 Colindale Avenue, London, NW9 5EQ. Conflicts of Interest: DB has received a research grant from Gilead. WLI has received speaker and consultancy fees from Roche, Janssen Cilag, Gilead Sciences and Novartis, educational grants from Boehringer Ingelheim, Merck Sharp and Dohme and Gilead Sciences, and research grant support from GlaxoSmithKline, Pfizer, Gilead Sciences, Janssen Cilag, Abbvie and Bristol-Myers Squibb. The remaining authors have no conflicts of interest. Author Contributions: WLI lead this work. DAS, DB and JS performed the analysis and wrote the first draft of the manuscript. This work was supervised by JMc, EB and WLI. DB, JM, CFM, JB, ET, AF, EAC, MAA, AB, EH performed viral sequencing and contributed analysis tools. WLI, JM and JB collected clinical data. DAS, DB, JM, CFM, DB, JS, ET, AF, EAC, MAA, AB, EH, JB, BH, PT, JMc, EB, WLI provided significant edits and feedback on the manuscript.

## Abstract

Sustained viral response (SVR) rates to first-line Direct Acting Antiviral (DAA) therapy for hepatitis C virus (HCV) infection routinely exceed 95%. However, a small number of patients require retreatment. Sofosbuvir, velpatasvir and voxilaprevir (SOF/VEL/VOX) is a potent DAA combination primarily used for the retreatment of patients failed by first line DAA therapies. Here we evaluate retreatment outcomes and the effects of resistance associated substitutions (RAS) in a real-world cohort, including the largest number of genotype (GT)3 infected patients, to date. 144 patients from the UK were retreated with SOF/VEL/VOX following virologic failure with first-line DAA treatment regimens. Full-length HCV genome, next-generation sequencing was performed prior to retreatment with SOF/VEL/VOX. HCV subtypes were assigned and RAS relevant to each genotype were identified (15% read cut-off). GT1a and GT3a were the two most common subtypes in the cohort, each making up 38% (GT1a n=55, GT3a n=54) of the cohort. 40% (n=58) of patients had liver cirrhosis of whom 7% (n=4) were decompensated, 10% (n=14) had hepatocellular carcinoma (HCC) and 8% (n=12) had received a liver transplant prior to retreatment. The overall re-treatment SVR12 rate was 90% (129/144). On univariate analysis, GT3 infection (50/62; SVR=81%, p=0.009), cirrhosis (47/58; SVR=81%, p=0.01) and prior treatment with SOF/VEL(12/17; SVR=71%, p=0.02) or SOF + DCV (14/19; SVR=74%, p=0.012) were all significantly associated with retreatment failure, but existence of pre retreatment RAS was not when the genotype of the virus is taken into account. The lower SVR rates achieved in patients retreated with SOF/VEL/VOX for patients with GT3 infection, cirrhosis and prior treatment with SOF/VEL or SOF/DCV has important implications for both patients and HCV elimination strategies.

## Introduction

Sustained Virological Response (SVR) rates for patients chronically infected with hepatitis C virus (HCV) and treated with Direct Acting Antiviral (DAA) therapies in clinical trials and real-world cohorts are often >95% (1–3). Treatment failure is associated with multiple host and viral factors including advanced fibrosis or cirrhosis and the presence of resistance associated substitutions (RAS) in HCV-encoded proteins that are targeted by DAA (4). HCV subtype is also known to have an effect on treatment outcomes; GT4r, GT3b and GT1l have each shown reduced susceptibility to NS5A inhibitor-containing regimens such as sofosbuvir and ledipasvir (SOF/LDV) or sofosbuvir and velpatasvir (SOF/VEL) (5–7). This means there is a small percentage of HCV-infected patients who have been failed by first-line therapies and are therefore by definition “difficult to treat”. Additionally, retreatment of this “difficult to treat” patient population may be made more challenging, as treatment failure commonly is associated with the emergence of RAS, which in the case of RAS to the NS5A inhibitor class of DAAs can persist for years after treatment failure (8). For patients who have been failed by pan-genotypic regimens such as SOF/VEL and glecaprevir and pibrentasvir, (GLE/PIB) retreatment options are limited. Currently the European Association for the Study of the Liver (EASL) and American Association for the Study of Liver Disease (AASLD), only recommend a combination of sofosbuvir, velpatasvir and voxilaprevir (SOF/VEL/VOX) for these patients(9,10). However GLE/PIB + SOF is effective in small clinical trials(11) and both AASLD and EASL consider it a valid alternative if patients have failed multiple rounds of therapy.

Retreatment of patients previously failed by predominantly first generation NS5A inhibitor-containing DAA therapy with SOF/VEL/VOX has been evaluated in both the POLARIS-1 and POLARIS-4 phase-II and III studies (12), with SVR rates in excess of 95%. In these trials most patients retreated with SOF/VEL/VOX (228/445; 51%) and (132/445; 29%) were infected with HCV GT1 and GT3 respectively, 46% of the patients had cirrhosis and 49-83% had detectable RAS before retreatment. Nevertheless, SOF/VEL/VOX showed a very high SVR rate in GT1 infected patients (222/228; 97% SVR); however, a slightly higher virologic failure rate in GT3 patients was observed (126/132; 95% SVR). A number of studies have evaluated the observed real-world outcomes of SOF/VEL/VOX retreatment, in both the US and Europe. The largest cohort (n=573) from the US, reported by Belperio, et al (13), showed lower SVR rates than in the POLARIS 1 and 4 studies for all genotypes (GT1: 429/473; 91% SVR, GT2: 18/20; 90% SVR, GT3: 42/46; 91% SVR, GT4: 12/12; 100% SVR). This study also showed that the SVR rate for those who had received SOF/VEL as a first-line therapy, was reduced for GTs 1-3 (GT1: 15/19; 79% SVR, GT2: 13/15; 87% SVR, GT3: 11/13; 85% SVR). In an Italian cohort (14), the SVR rate was 95% (162/169) with cirrhosis and hepatocellular carcinoma (HCC) being associated with treatment failure, however there was no significant difference between the SVR rates of patients with different genotypes (GT1: 98/101; 97% SVR, GT2: 17/17; 100% SVR, GT3: 33/36; 91% SVR, GT4: 14/15; 93% SVR). Finally, Llaneras, et al (15) reported an overall SVR rate of 95% (128/135) with 100% in patients with GT1 (82/82) and GT2 (7/7) infection, 80% (24/30) in GT3 and 93% (13/14) in GT4. SVR rates were significantly lower in patients with cirrhosis (89%, p = 0.05), or those with GT3 infection (80%, p <0.001), whilst patients with GT3 infection and cirrhosis had the lowest SVR rate (69%).

Despite these studies there remains limited data from the real-world setting, particularly in patients with GT3 infection and on the prevalence of post-treatment RAS amongst patients not achieving SVR with first line DAA treatment and the potential impact of these RAS on SOF/VEL/VOX retreatment outcomes. Data following unsuccessful therapy with pan-genotypic DAA regimens such as SOF/VEL or GLE/PIB retreated with SOF/VEL/VOX is particularly lacking and these data are urgently required to inform optimal retreatment strategies. In this paper we report the outcomes of 144 patients failed by first line DAA therapy and retreated with SOF/VEL/VOX. We then analyse the effect of clinical characteristics and RAS on SOF/VEL/VOX retreatment.

## Results

### Baseline characteristics of patients

Outcomes were available for 144 patients who were retreated with SOF/VEL/VOX. The mean age was 56 years (49-63 IQR) with 84% (n=121) of the cohort being male. Overall, 40% (n=58) had cirrhosis, with 7% (n=4) decompensated. 10% (n=14) had HCC and 8% (n=12) had received a liver transplant prior to retreatment (Table 1). GT1a and GT3a were the two most common subtypes, each making up 38% (GT1a n=55, GT3a n=54) of the cohort, 17 other subtypes were identified from genotypes 1,2,3,4 and 6, with GT1b, GT4r and GT3b making up 6% (n=9),4% (n=6), and 3% (n=5) of the cohort respectively (Figure 1).

**Table 1:**
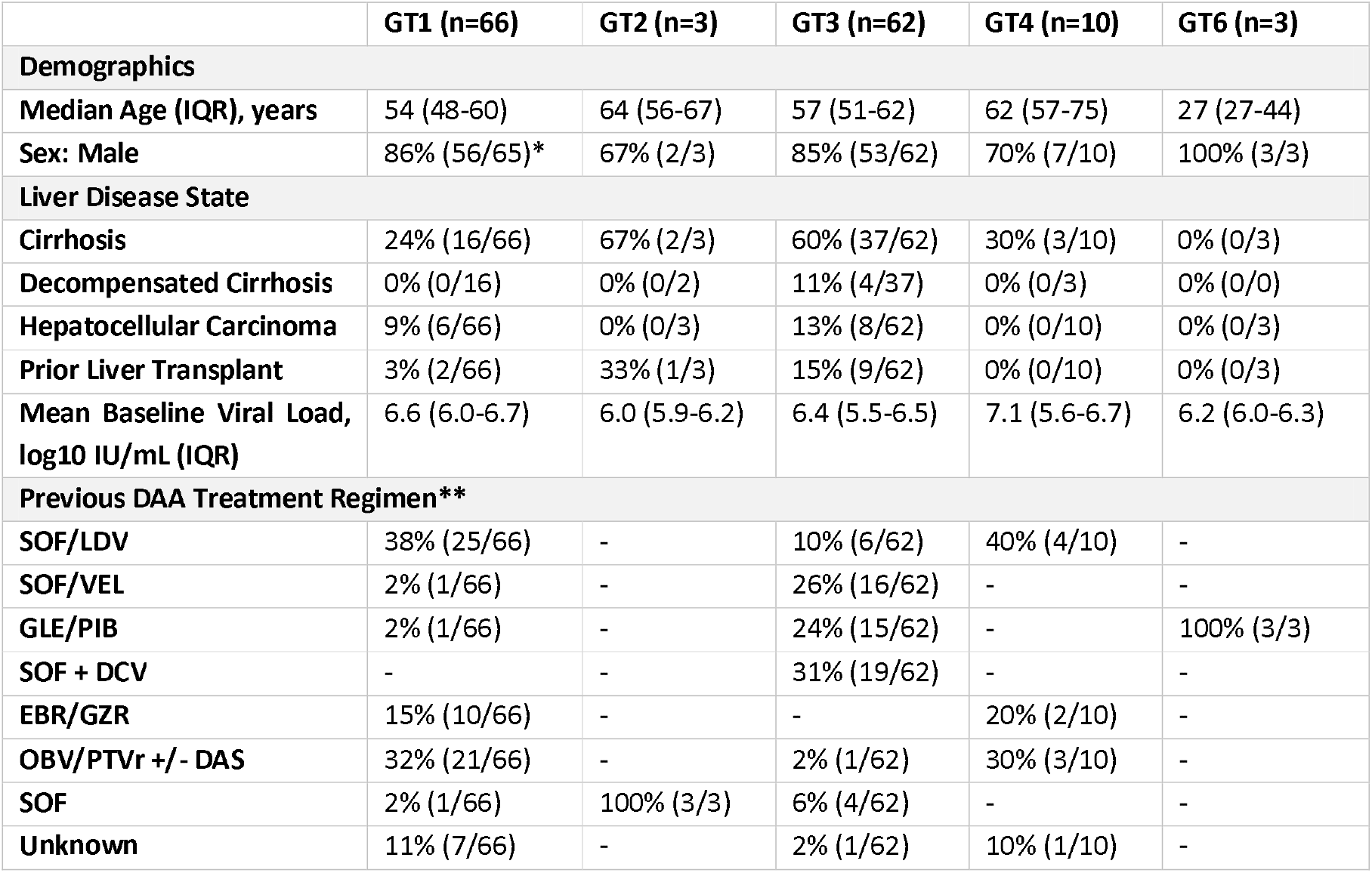
Clinical characteristics of 144 patients prior to retreatment with SOF/VEL/VOX split by genotype. Daclatasvir (DCV), Sofosbuvir (SOF), Glecaprevir (GLE), Pibrentasvir (PIB), Grazoprevir (GZR), Elbasvir (EBR), Ledipasvir (LDV), Paritaprevir (PTV), Ombitasvir (OBV) Ritonavir (r), Dasabuvir (DAS), Velpatasvir (VEL). *Data missing for one patient. **Full breakdown of previous treatment regimens including ribavirin and interferon use are in Supplementary Table 1

|  | GT1 (n=66) | GT2 (n=3) | GT3 (n=62) | GT4 (n=10) | GT6 (n=3) |
| --- | --- | --- | --- | --- | --- |
| **Demographics** |  |  |  |  |  |
| **Median Age (IQR), years** | 54 (48-60) | 64 (56-67) | 57 (51-62) | 62 (57-75) | 27 (27-44) |
| **Sex: Male** | 86% (56/65)* | 67% (2/3) | 85% (53/62) | 70% (7/10) | 100% (3/3) |
| **Liver Disease State** |  |  |  |  |  |
| **Cirrhosis** | 24% (16/66) | 67% (2/3) | 60% (37/62) | 30% (3/10) | 0% (0/3) |
| **Decompensated Cirrhosis** | 0% (0/16) | 0% (0/2) | 11% (4/37) | 0% (0/3) | 0% (0/0) |
| **Hepatocellular Carcinoma** | 9% (6/66) | 0% (0/3) | 13% (8/62) | 0% (0/10) | 0% (0/3) |
| **Prior Liver Transplant** | 3% (2/66) | 33% (1/3) | 15% (9/62) | 0% (0/10) | 0% (0/3) |
| **Mean Baseline Viral Load, log<sub>10</sub> IU/mL (IQR)** | 6.6 (6.0-6.7) | 6.0 (5.9-6.2) | 6.4 (5.5-6.5) | 7.1 (5.6-6.7) | 6.2 (6.0-6.3) |
| **Previous DAA Treatment Regimen\*\*** |  |  |  |  |  |
| **SOF/LDV** | 38% (25/66) | - | 10% (6/62) | 40% (4/10) | - |
| **SOF/VEL** | 2% (1/66) | - | 26% (16/62) | - | - |
| **GLE/PIB** | 2% (1/66) | - | 24% (15/62) | - | 100% (3/3) |
| **SOF + DCV** | - | - | 31% (19/62) | - | - |
| **EBR/GZR** | 15% (10/66) | - | - | 20% (2/10) | - |
| **OBV/PTVr +/- DAS** | 32% (21/66) | - | 2% (1/62) | 30% (3/10) | - |
| **SOF** | 2% (1/66) | 100% (3/3) | 6% (4/62) | - | - |
| **Unknown** | 11% (7/66) | - | 2% (1/62) | 10% (1/10) | - |

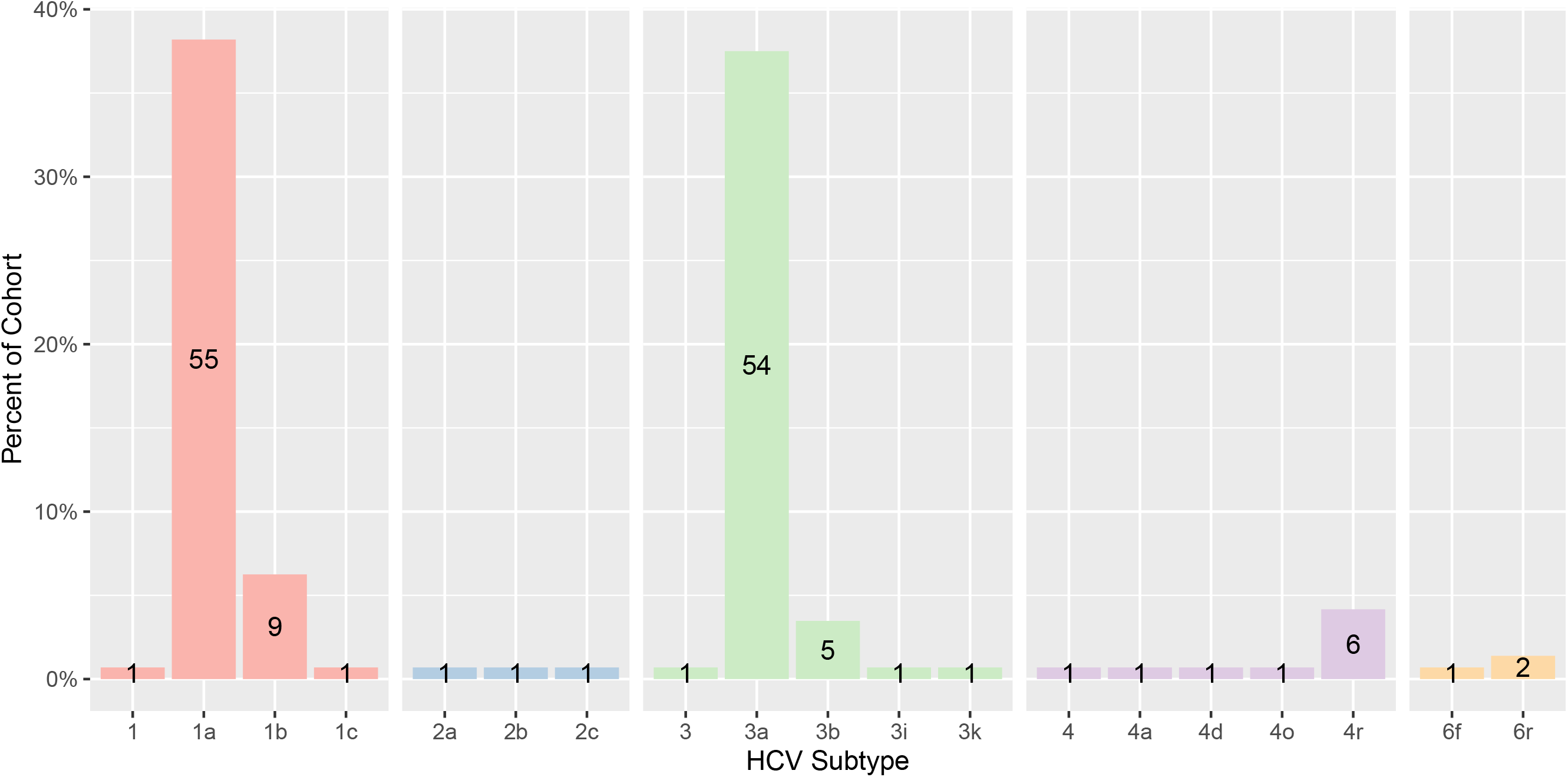

### Effectiveness of retreatment with SOF/VEL/VOX

The overall SVR12 rate for patients retreated with SOF/VEL/VOX was 91% (Figure 2). The majority of patients who did not achieve an SVR (n=15) experienced a post treatment relapse (n=10). One patient experienced an on-treatment breakthrough (patient’s HCV RNA is undetectable during treatment and then becomes detectable again during treatment.) and two were non-responders (patients who never achieve undetectable HCV RNA). Relapse/breakthrough/non-response was not reported by the treating clinician for the remaining two patients.

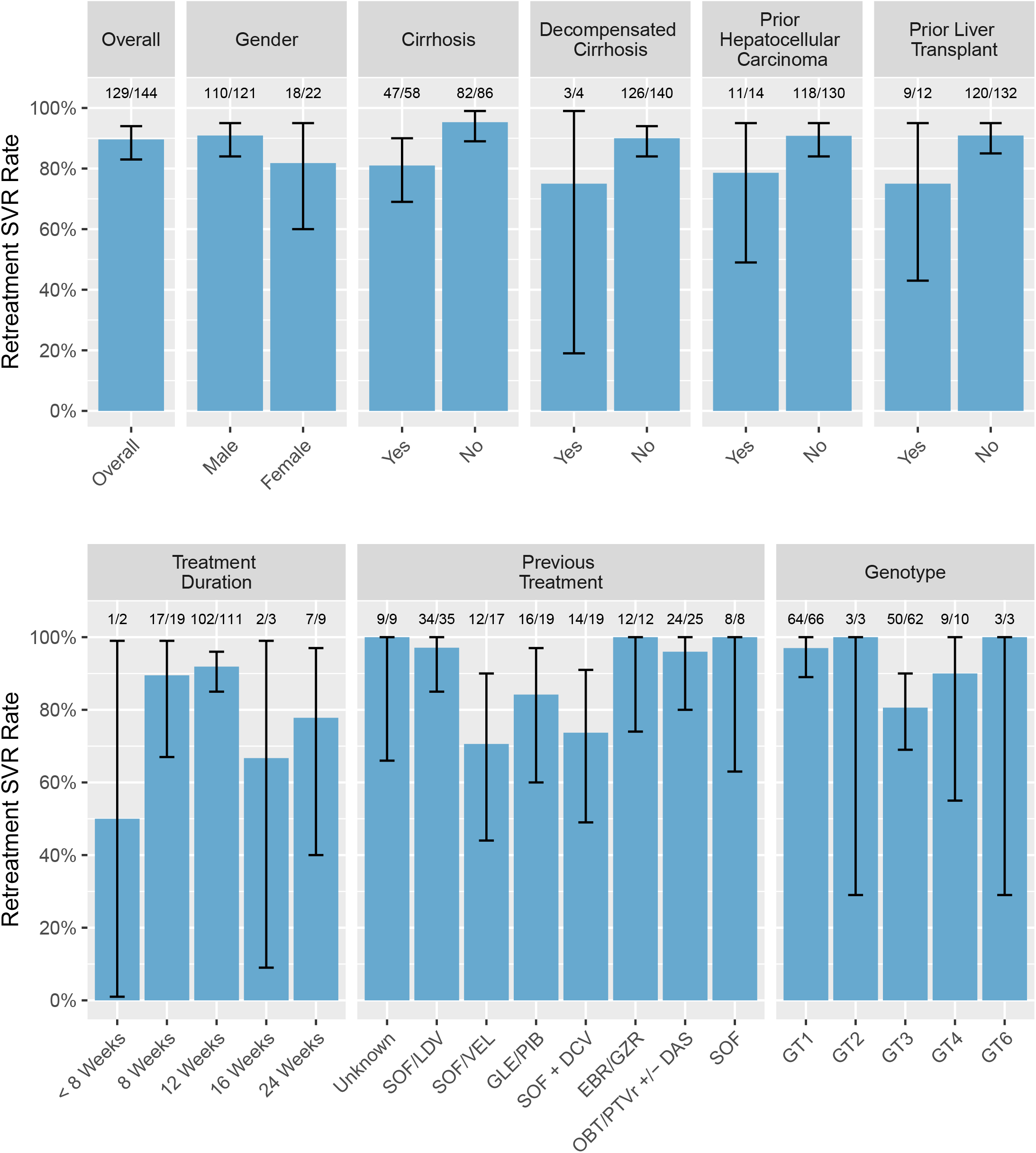

Evaluating the factors associated with outcome using univariable logistic regression, GT3 infection (52/64; SVR=81%, p=0.009) and cirrhosis (47/58; SVR=81%, p=0.01) were both significantly associated with treatment outcome (Figure 3). Patients whose initial regimen was either SOF/VEL or SOF + DAC were also less likely to achieve SVR on retreatment (12/17, 71% =0.02 and 14/19, 74%, p=0.012 respectively). Patients with GT3 infection, cirrhosis and prior treatment with SOF/VEL had the lowest SVR rate of 42% (3/7). However, one of these patients only received 4 weeks of SOF/VEL/VOX (Supplementary Table 2). Multivariable analysis using logistic regression with SVR as the response variable and GT, presence/absence of cirrhosis and previous treatment regimen included as explanatory variables, revealed that prior treatment with SOF/VEL had the largest effect on SVR and remained the only variable significantly associated with outcome (p=0.03) (Supplementary Figure 1). However, with the exception of one GT1 patient previously treated with SOF/VEL, all patients with prior SOF/VEL or SOF + DCV treatment had GT3 infection which confounded this result (Table 1). When GT3-infected patients were analysed separately, cirrhosis and prior treatment regimen were no longer significantly associated with outcome, in both the univariable and multivariable analysis.

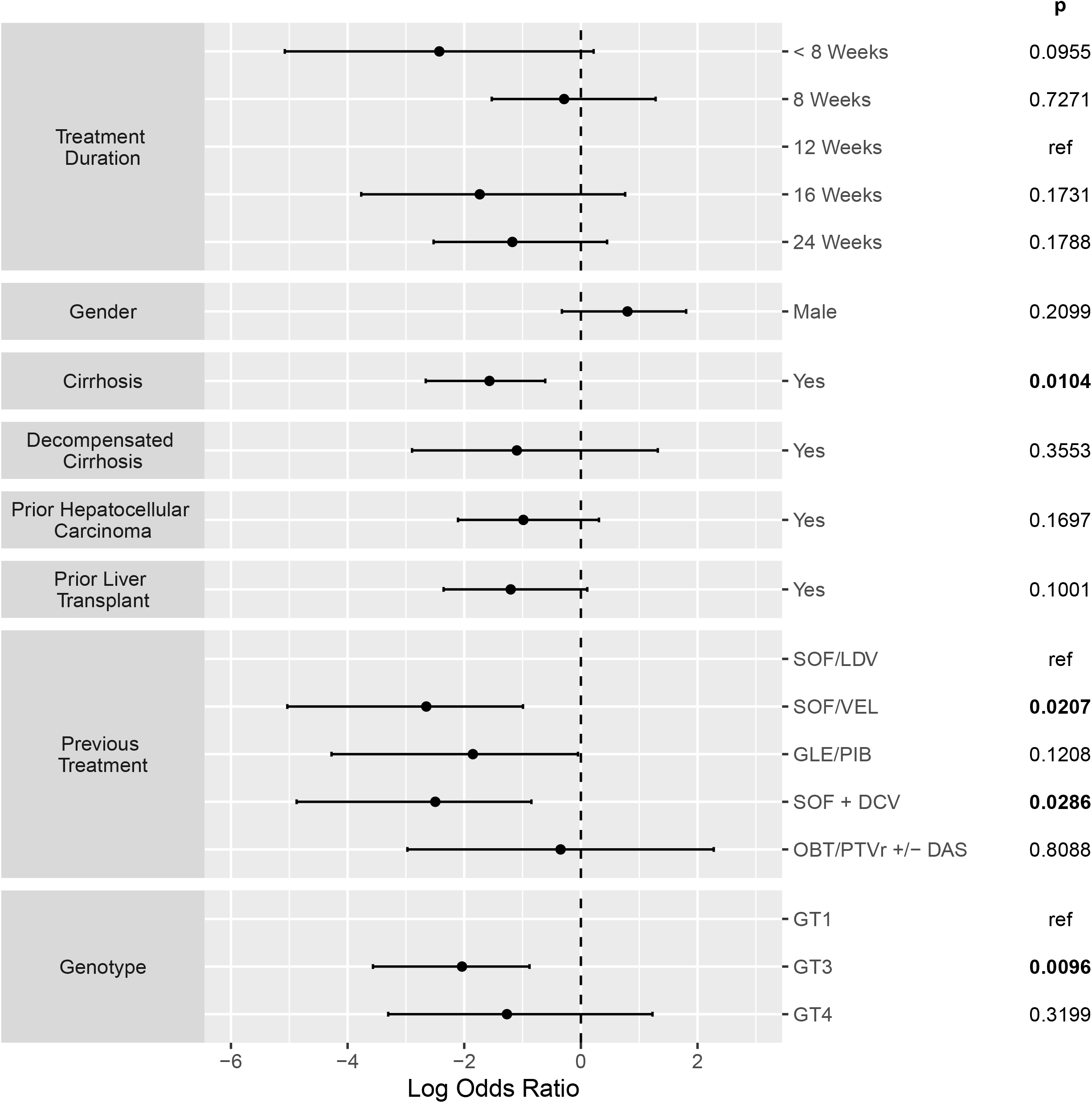

### Effect of resistance-associated substitutions on retreatment outcome

Among the 144 patients retreated with SOF/VEL/VOX and with available outcomes, 70% (101/144) had detectable RAS prior to retreatment. 16% had NS3 inhibitor RAS, 51% NS5A inhibitor RAS and 34% SOF RAS. Of the viral sequences in 15 patients failed by SOF/VEL/VOX retreatment, 12/15 (80%) had detectable RAS prior to retreatment compared to 89/129 (68%) in those subsequently achieving SVR. However, despite the high prevalence of RAS, the presence of a RAS in a particular protein or combination of proteins was not significantly associated with outcome (Table 2). When each RAS was considered independently using logistic regression, A30K and Y93H RAS in NS5A were associated with retreatment failure in the whole cohort (A30K p=0.02, Y93H p=0.01) (Table 3). The A30K RAS is unique to GT3a (it is wild-type in GT3b) and the Y93H substitution was much more common in GT3 patients compared to other genotypes (Supplementary Figure 2). When the analysis is limited to GT3 patients, the SVR rates for patients with these RAS were still reduced (for A30K SVR, 71% and Y93H SVR, 78%) but this was no longer statistically significant (A30K p=0.2, Y93H p=0.3). In addition, when genotype is added as a cofactor to the logistic regression Y93H is no longer associated with outcome. Whilst individually the A30K and Y93H RAS are common, the combination of A30K + Y93H is found rarely and is not associated with treatment outcome in this cohort (Supplementary Figure 3). No other RAS was significantly associated with outcome within the whole cohort or within a specific genotype of patients.

**Table 2:** Prevalence of pre-retreatment RAS and SVR rates for patients retreated with SOF/VEL/VOX. Logistic regression was used to test the association between presence of RAS in the NS3, NS5A and NS5B proteins with SVR.

| Presence of RAS | Prevalence % (n) | SVR Rate % (n) | p | log Odds Ratio | Standard Error |
| --- | --- | --- | --- | --- | --- |
| **No RAS** | 30% (43/144) | 93% (40/43) | NA | NA | NA |
| **NS3** | 16% (23/144) | 100% (23/23) | 0.9 | 15.9 | 2062 |
| **NS5A** | 51% (74/144) | 86% (64/74) | 0.3 | -0.7 | 0.8 |
| **NS5B** | 34% (49/144) | 84% (41/49) | 0.5 | -0.5 | 0.96 |
| **NS3 + NS5A** | 8% (12/144) | 100% (12/12) | 0.9 | 15.9 | 1882 |
| **NS3 + NS5B** | 0% (0/144) | NA | NA | NA | NA |
| **NS5A + NS5B** | 22% (31/144) | 80% (25/31) | 0.1 | -1.1 | 0.75 |
| **NS3 + NS5A + NS5B** | 1% (1/144) | 100% (1/1) | 0.9 | 15.9 | 6522 |

**Table 3:**
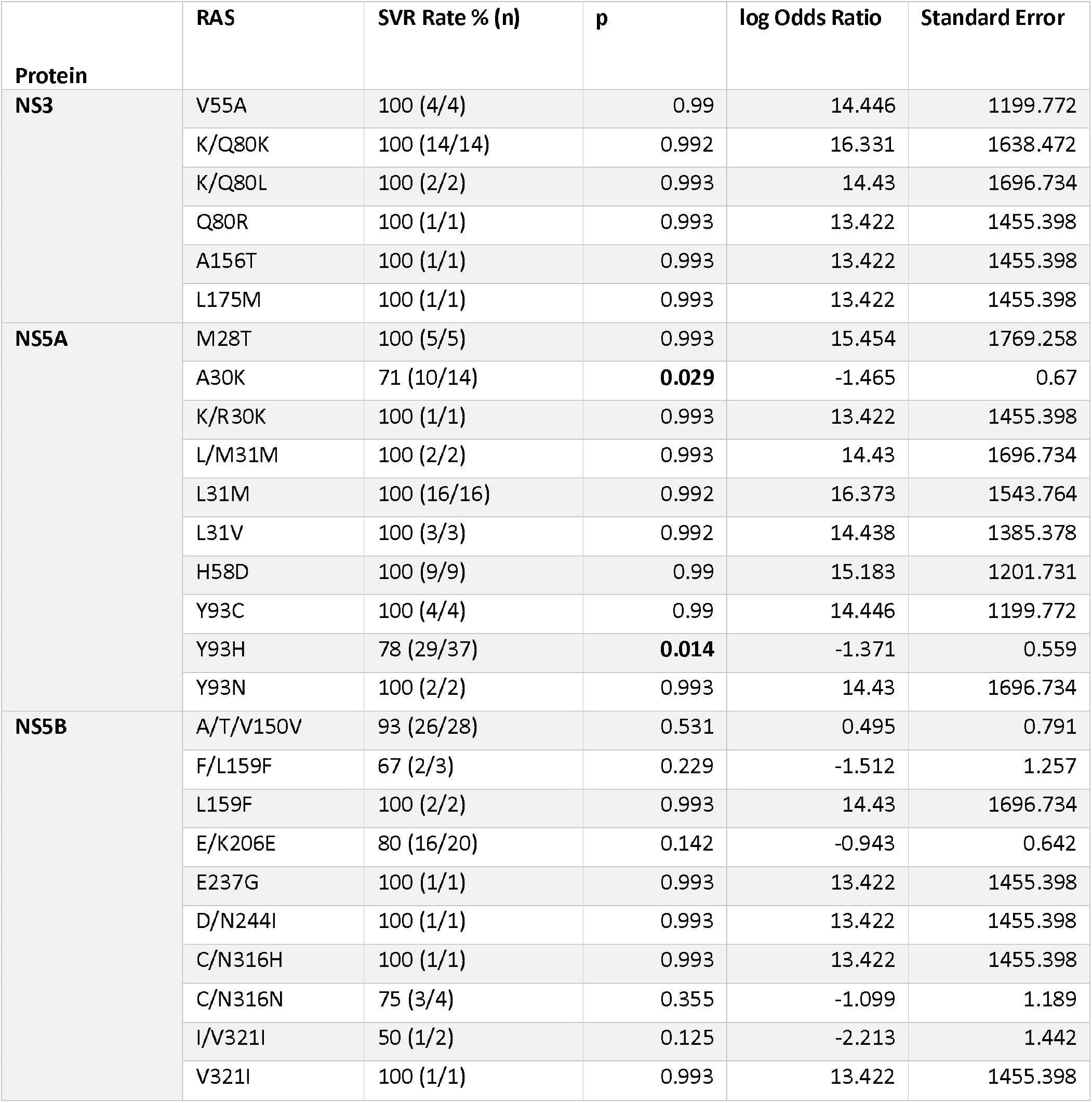
SVR rates for patients retreated with SOF/VEL/VOX with individual pre retreatment RAS. Logistic regression was used to test the association between presence of RAS with outcome in the whole cohort.

## Discussion

This cohort represents both the largest reported cohort of patients retreated with SOF/VEL/VOX in the UK, and the largest cohort of GT3-infected patients retreated with SOF/VEL/VOX. Our results show that retreatment of patients with SOF/VEL/VOX who have been failed by first-line therapy is very effective for GT1. However, patients with GT3 infection, cirrhosis or prior treatment with SOF/VEL or SOF/DCV have significantly lower SVR rates with SOF/VEL/VOX retreatment.

Our data shows that GT3-infected patients achieved lower SVR rates to SOF/VEL/VOX retreatment; the lowest SVR12 rates (42%) were observed in patients with GT3 infection, cirrhosis and prior SOF/VEL exposure. This is consistent with the data from Llaneras et al (15), which also showed a SVR rate of 81% for GT3-infected patients and 69% for GT3-infected patients with cirrhosis. Belperio, et al (13) have shown that GT1-infected patients with prior SOF/VEL exposure had a reduced SVR rate of 79% (15/19), yet a GT3-infected group with prior SOF/VEL exposure had a SVR rate of 85% (11/13) which is greater than the 75%(12/16) that was observed in the GT3-infected patients in our study but, these numbers remain small.

The overall prevalence of RAS was very high in this cohort and the presence of the NS5A inhibitor RAS Y93H was significantly associated with treatment failure across the whole cohort. However, when genotype is included in the model used to test association, Y93H is no longer significantly associated with outcome. This data suggests that viral genotype is the more important viral factor for SOF/VEL/VOX retreatment outcome than the presence of RAS. However, PIB has been shown to be more effective than other NS5A inhibitors against viruses harbouring the Y93H RAS in-vitro (16). This could mean that GLE/PIB would be more effective against virus strains harbouring the Y93H RAS. Thus, RAS testing to guide retreatment therapy could be an effective way to increase retreatment SVR rates.

At present SOF/VEL/VOX is the only recommended retreatment regimen for patients previously failed by a NS5A inhibitor containing regimen in England. These data suggest that whilst SOF/VEL/VOX may be a preferred regimen for non-GT3 infected patients, an alternative retreatment regimen for GT3-infected patients should also be considered, particularly those with cirrhosis and previous exposure to a DAA regimen containing VEL or DCV. One current alternative to SOF/VEL/VOX for these patients is the off-label use of GLE/PIB + SOF +/-RBV. This combination was effective in a small retreatment trial (11) and could represent an effective retreatment option for GT3 patients with prior exposure to SOF/VEL. Further options include adopting a resistance guided approach to treatment, increasing the SOF/VEL/VOX treatment duration or adding ribavirin.

In summary, our study shows that retreatment outcomes for patients failed by first-line DAA therapy is very successful for non-GT3-infected patients. However, for GT3-infected patients, particularly those with cirrhosis and failed by initial SOF/VEL treatment, SVR rates were significantly lower and alternative retreatment regimens such as GLE/PIB + SOF should be considered.

## Methods

### Patients and Samples

This study included 215 individuals from across England, United Kingdom who did not achieve a SVR with previous interferon-free DAA treatment. Blood samples were taken prior to retreatment and samples sent to Oxford and Glasgow for HCV whole genome sequencing (WGS). Clinical, demographic and treatment outcome data were collected from patients, who were enrolled in HCV Research UK (n=37), following informed consent and ethical approval (17). For patients not enrolled in HCV Research UK, whole genome HCV sequences and anonymised clinical data were provided by Public Health England (PHE), collected as part of the Virus Reference Department’s ‘s HCV antiviral resistance testing service. This provides a clinical service for National Health Service Trusts, which send patients’ blood samples for HCV genotyping and resistance testing by WGS and receive results in real time to inform clinical management. Approval for the use of data from this service was granted under Regulation 3 of the Health Service (Control of Patient Information) Regulations 2002. For retreatment, SVR was defined a minimum of 12 weeks after the end of treatment.

### Whole Genome Sequencing of HCV

Whole HCV genome sequencing was conducted at three sites to define HCV subtype and identify RAS: (i) The MRC-University of Glasgow Centre for Virus Research (CVR), (ii) The Peter Medawar Building for Pathogen Research, University of Oxford, and (iii) The Virus Reference Department, National Infection Service, Public Health England. These sites have previously collaborated to ensure uniform standards for HCV sequencing (18). Briefly, RNA was isolated from patient plasma samples and reverse transcribed to produce cDNA. An Illumina sequencing library was generated and enriched for HCV viral sequences using specific oligonucleotide probes. Illumina sequencing reads were then processed and mapped to the closest HCV reference genome. Precise details of how the sequencing was performed at each site is described in the supplementary materials.

### Resistance Associated Substitution Calling using HCV-GLUE

HCV subtypes were assigned and NS3, NS5A and NS5B RAS relevant to each genotype were identified (15% of reads cut off) using HCV-GLUE and RAS definitions provided by PHE (19) (Supplementary Table 3). HCV-GLUE is a resource created as part of the STOP-HCV consortium based on the GLUE (Genes Linked by Underlying Evolution) platform (20). HCV-GLUE uses the ICCT HCV reference sequences and collates publicly available HCV sequences, to construct phylogenies using RAxML to assign sequences to HCV clades. It also contains a database of HCV RAS created by PHE (19) and this is used to identify RAS relevant to the HCV genotype and subtype (21).

### Statistical Analysis

All statistical analysis was performed using custom code and R version 3.6.2. Logistic regression was used for analysis of association between categorical variables.

## Data Availability

Data can be made avalible on requast.

## Acknowledgments

The following clinicians contributed to data collection for this project:

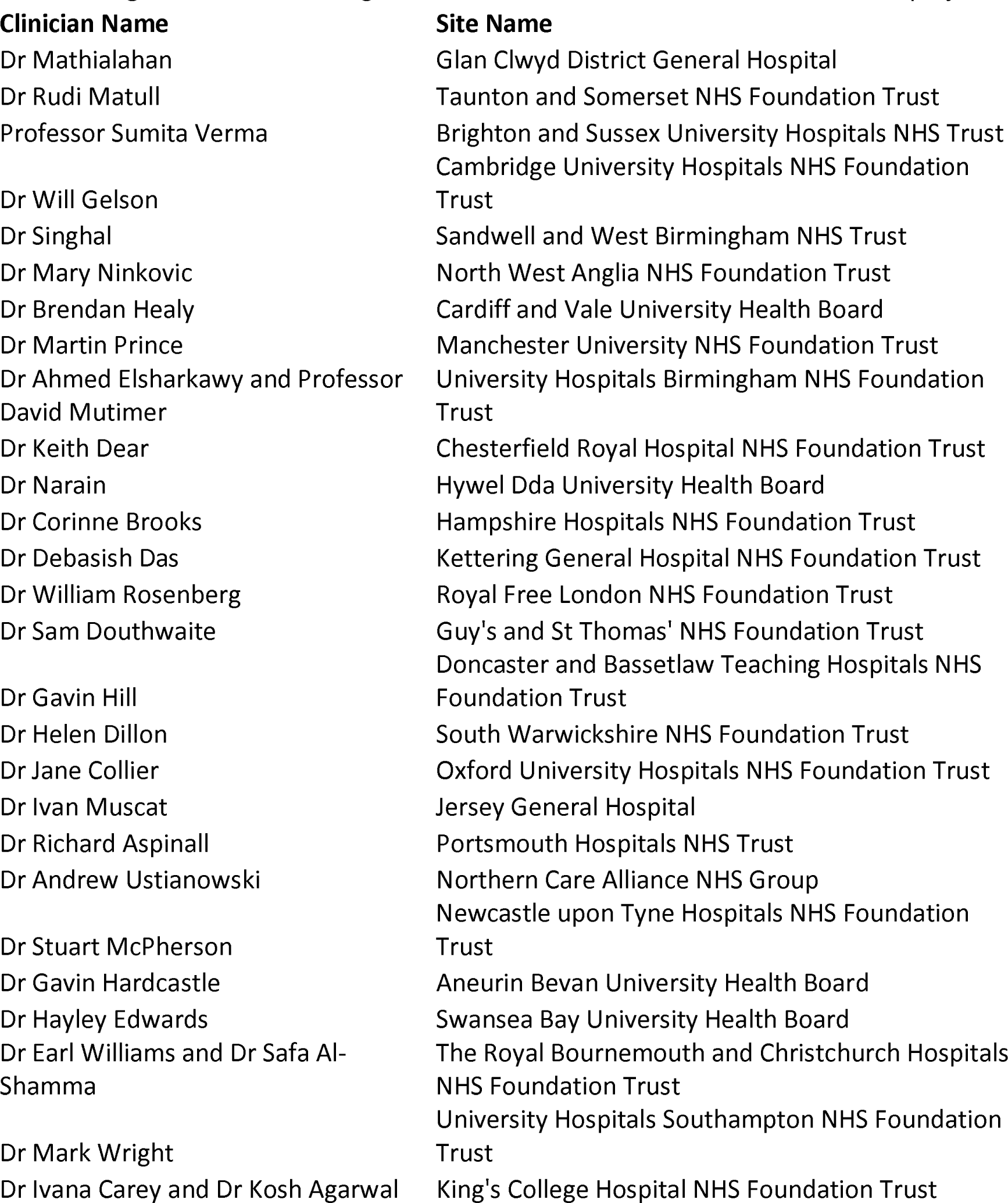
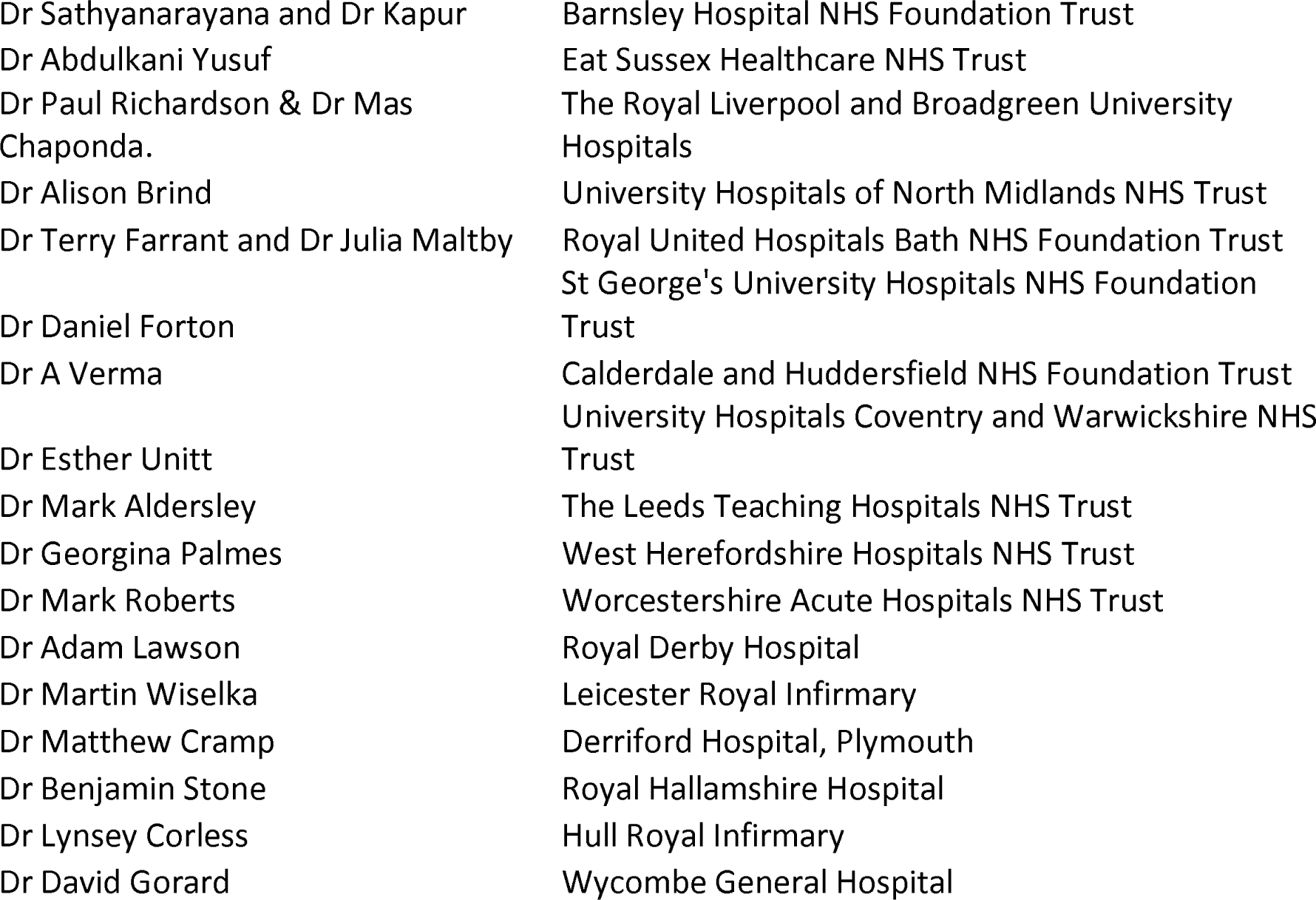

## Abbreviations

SVR: Sustained Viral Response
HCV: Hepatitis C Virus
DAA: Direct Acting Antiviral
RAS: Resistance Associated Substitution
NS: Non-Structural
SOF: Sofosbuvir
LDV: Ledipasvir
VEL: Velpatasvir
GLE: Glecaprevir
PIB: Pibrentasvir
EASL: European Association for the Study of the Liver
AASLD: American Association for the Study of Liver Disease
VOX: Voxilaprevir
GT: Genotype
HCC: Hepatocellular Carcinoma
DCV: Daclatasvir
RBV: Ribavirin
PHE: Public Health England
WGS: Whole Genome Sequencing
GLUE: Genes Linked by Underlying Evolution

