## Supplementary for "Real World SOF/VEL/VOX Retreatment Outcomes and Viral Resistance Analysis for HCV Patients with Prior Failure to DAAs"

### Supplementary Materials

#### Methods

##### Sequencing at MRC-University of Glasgow Centre for Virus Research

RNA was isolated from 200 μL of plasma using the RNAdvance Blood extraction kit (Beckman Coulter, Brea, CA, United States) and collected in 27 μL of water. Following conversion of RNA to double-stranded DNA, libraries were prepared for Illumina sequencing using the KAPA DNA LTP Library Preparation Kit (Roche, Basel, Switzerland), and NEBNext Multiplex Oligos for Illumina (New England Biolabs, Ipswich, MA, United States). Libraries were quantified using Qubit dsDNA HS Assay Kit (Invitrogen, Carlsbad, CA, United States) and size distribution assessed using Agilent TapeStation with D1K High Sensitivity Kit (Agilent, Santa Clara, CA, United States); libraries were normalised according to viral load and mass. A 500 ng aliquot of the pooled library was enriched using SeqCap EZ Developer Probes (Roche), following the manufacturer’s protocol. Following a 14 cycle post-enrichment PCR, the cleaned pool was sequenced with 151-base paired-end reads on a NextSeq cartridge (Illumina, San Diego, CA, United States). Reads were trimmed and filtered using TrimGalore^23^ with a quality threshold of 30 and minimum read length of 75 b. The most appropriate HCV reference sequence was identified via a k-mer-based approach, using k-mers unique to each genotype. BAM files were generated by mapping against the best-matching HCV reference using Tanoti (https://bioinformatics.cvr.ac.uk/software/tanoti/)^22^.

##### Sequencing at National Infection Service, Public Health England

The PHE library preparation protocol is the laboratory component of a pipeline aimed at clinical use; a manuscript describing the full pipeline is in preparation. RNA was extracted from 350 μL of plasma using the NUCLISENS easyMAG system (bioMerieux). Total eluates were subjected to Turbo DNAse treatment (Thermo Fisher, Waltham, MA, United States) followed by library preparation using KAPA RNA HyperPrep kit (Roche). Libraries were pooled based upon DNA concentration and HCV quantity, assessed using the Quant-iT kit on the Glomax platform (Promega, Madison, WI, United States) and the Qiagen QuantiTect kit with primers and probes from Davalieva et al.^24^. Pools were enriched by hybridisation to a biotinylated probe set (Integrated DNA Technologies, described by Bonsall et al^25^) followed by further PCR cycles depending upon HCV quantity. The two pools were pooled, again by concentration and HCV quantity. The final pool was quantified using the KAPA SYBR FASTA qPCR kit (Roche) on a PRISM 7500 (Applied BioSystems, Foster City, CA, United States) before being sequenced on a MiSeq using Reagent kit v2 (Illumina). Human reads were filtered out from trimmed FASTQ files using SMALT^26^, remaining reads were then assembled using VICUNA de novo assembly ^27^. Contigs were matched to HCV reference genomes using BLAST^28^ and gaps filled using LASTZ^29^ to generate a draft assembly. Reads were then mapped to the draft assembly with BWA^30^.

##### Sequencing at Peter Medawar Building for Pathogen Research University of Oxford

RNA was extracted from 500μl of plasma using the NucliSENS®easyMAGsystem(bioMe ́rieux) into 30 μl of water, of which 5 μl was processed with the NEBNext® UltraTMDirectional RNA Library Prep Kit for Illumina® (New England Biolabs) with previously published modifications to the manufacturer’s protocol^31^. A 500 ng aliquot of the pooled library was enriched using the xGen® Lockdown® protocol from IDT (Rapid Protocol for DNA Probe Hybridisation and Target Capture Using an Illumina TruSeq® Library (v1.0), Integrated DNA Technologies) with a comprehensive panel of HCV-specific 120 nt DNA oligonucleotide probes (IDT), designed using a previously published algorithm^25^. The enriched library was sequenced using Illumina MiSeq v2 chemistry to produce paired 150 b reads. Reads were demultiplexed and 150 b paired-end reads were trimmed using QUASR v7.01^32^ then adaptor sequences removed using Cutadapt v1.7.1^33^ reads were discarded if less than 50p long. Reads were mapped to human reference sequence using Bowtie 2.2.4^34^ and host-derived sequences were removed. The remaining reads were then mapped against a database containing all 165 ICTV (International Committee of the Taxonomy of Viruses)^35^ HCV genomes using BWA mem v0.7.10^36^ and STAMPY v1.0.23^30^. An appropriate reference sequence was selected from this database and reads which conform to the majority population were then used for de novo assembly using Vicuna v1.3^27^ and V-FAT v1.0^37^. All viral reads were then mapped back to this assembly using Mosaik v2.2.28^38^ to generate a BAM file containing viral reads. Further detail of the techniques used can be found in Bonsall et al, 2015^25^.

### Supplementary Tables

|  | GT1 (n=66) | GT2 (n=3) | GT3 (n=62) | GT4 (n=10) | GT6 (n=3) |
| --- | --- | --- | --- | --- | --- |
| DCV + SOF | - | - | 3% (2/62) | - | - |
| DCV + SOF + IFN + RBV | - | - | 8% (5/62) | - | - |
| DCV + SOF + RBV | - | - | 19% (12/62) | - | - |
| EBR/GZR | 12% (8/66) | - | - | 10% (1/10) | - |
| EBR/GZR + RBV | 3% (2/66) | - | - | 10% (1/10) | - |
| GLE/PIB | 2% (1/66) | - | 23% (14/62) | - | 100% (3/3) |
| GLE/PIB + RBV | - | - | 2% (1/62) | - | - |
| OBT/PTVr | 3% (2/66) | - | - | - | - |
| OBT/PTVr + RBV | - | - | - | 10% (1/10) | - |
| OBT/PTVr + DAS | 15% (10/66) | - | 2% (1/62) | - | - |
| OBT/PTVr + DAS + RBV | 12% (8/66) | - | - | 20% (2/10) | - |
| OBT/PTVr + DAS + IFN + RBV | 2% (1/66) | - | - | - | - |
| SOF | - | 33% (1/3) | 2% (1/62) | - | - |
| SOF + RBV | 2% (1/66) | 67% (2/3) | - | - | - |
| SOF + IFN + RBV | - | - | 5% (3/62) | - | - |
| SOF/LDV | 29% (19/66) | - | 5% (3/62) | 30% (3/10) | - |
| SOF/LDV + RBV | 8% (5/66) | - | 5% (3/62) | 10% (1/10) | - |
| SOF/LDV + IFN + RBV | 2% (1/66) | - | - | - | - |
| SOF/VEL | 2% (1/66) | - | 23% (14/62) | - | - |
| SOF/VEL + RBV | - | - | 2% (1/62) | - | - |
| SOF/VEL + IFN | - | - | 2% (1/62) | - | - |
| Unknown | 11% (7/66) | - | 2% (1/62) | 10% (1/10) | - |

**Supplementary Table 1: Full breakdown of patients previous treatment regimens:** Daclatasvir (DCV), Sofosbuvir (SOF), Glecaprevir (GLE), Pibrentasvir (PIB), Grazoprevir (GZR), Elbasvir (EBR), Ledipasvir (LDV), Paritaprevir (PTV), Ombitasvir (OBV) Ritonavir (r), Dasabuvir (DAS), Velpatasvir (VEL), Ribavirin (RBV), Interferon (INF).

| Age | Gender | HCV Subtype | Treatment Length | Outcome | Presence of Cirrhosis | Decompensated Cirrhosis | Previous Hepatocellular Carcinoma | Previous Liver Transplant | Previous Treatment | Presence of RAS | Viral Load |
| --- | --- | --- | --- | --- | --- | --- | --- | --- | --- | --- | --- |
| 39 | Male | 3a | 12 Weeks | Relapse | Yes | No | No | No | GLE/PIB | NS5A + NS5B | 4146303 |
| 55 | Male | 3b | 12 Weeks | Relapse | Yes | No | No | Yes | SOF/VEL | No RAS | 3630780 |
| 53 | Male | 1b | 12 Weeks | Relapse | Yes | No | Yes | No | SOF/VEL | NS5A + NS5B | 3311311 |
| 44 | Female | 3a | 8 Weeks | Non-Responder | No | No | No | No | SOF + DAC + RBV | NS5A | 2206 |
| 51 | Male | 3a | 12 Weeks | Relapse | Yes | No | No | No | DCV + SOF + RBV | NS5A + NS5B | 1170000 |
| 51 | Male | 3a | 12 Weeks | Relapse | No | No | No | No | DCV + SOF + RBV | NS5A + NS5B | 1170000 |
| 49 | Male | 3a | 4 Weeks | No SVR* | Yes | No | No | No | SOF/VEL | NS5B | NA |
| 62 | Male | 3a | 24 Weeks | Breakthrough | Yes | Yes | Yes | No | SOF/VEL | No RAS | NA |
| 53 | Male | 3a | 24 Weeks | Relapse | Yes | No | No | Yes | DCV + SOF | NS5A | NA |
| 61 | Male | 3a | 12 Weeks | Relapse | Yes | No | No | No | DCV + SOF + RBV | NS5A + NS5B | 1300000 |
| 57 | Male | 3a | 8 Weeks | Non-Responder | No | No | No | No | GLE/PIB | NS5A | NA |
| 59 | Female | 4r | 12 Weeks | Relapse | Yes | No | No | No | SOF/LDV | NS5B | 6527426 |
| 71 | Male | 3a | 12 Weeks | Relapse | Yes | No | No | Yes | SOF/VEL | NS5A + NS5B | 92349 |
| 77 | Female | 3k | 16 Weeks | Relapse | Yes | No | Yes | No | GLE/PIB | NS5A | 2997956 |
| 54 | Female | 1a | 12 Weeks | No SVR* | No | No | No | No | OBT/PTVr + DAS | No RAS | 3603908 |

**Supplementary Table 2: Clinical characteristics of the 15 individual patients who were failed by retreatment.** Daclatasvir (DCV), Sofosbuvir (SOF), Glecaprevir (GLE), Pibrentasvir (PIB), Grazoprevir (GZR), Elbasvir (EBR), Ledipasvir (LDV), Paritaprevir (PTV), Ombitasvir (OBV) Ritonavir (r), Dasabuvir (DAS), Velpatasvir (VEL). * Reason not recorded.

| Drug | Genotypes | RAS |
| --- | --- | --- |
| voxilaprevir | 1a | 36G |
|  | 3a, 6a | 41K |
|  | 1a, 4a, 6a | 41R |
|  | 1a | 43S |
|  | 2a | 43V |
|  | 1a | 55A |
|  | 6a | 56H |
|  | 1a, 3a | 80K |
|  | 1a | 80L |
|  | 3a | 80R |
|  | 1b | 122D |
|  | 1a, 1b | 155W |
|  | 1a, 2a, 4a | 156L |
|  | 1b | 156S |
|  | 1a, 1b, 2a, 3a, 4a | 156T |
|  | 1b, 2a, 3a, 4a | 156V |
|  | 5a, 6a | 168A |
|  | 4a, 5a | 168E |
|  | 1a | 168F |
|  | 5a, 6a | 168H |
|  | 1a | 168I |
|  | 1a, 5a | 168K |
|  | 1a | 168L |
|  | 1a, 5a | 168R |
|  | 1a, 4a | 168T |
|  | 4a | 168V |
|  | 1b, 5a | 168Y |
|  | 1b | 170A |
|  | 3a | 175M |
| velpatasvir | 1a | 24R |
|  | 2a, 6a | 28S |
|  | 1a | 28T |
|  | 1a | 30E |
|  | 1a, 3a | 30K |
|  | 4d | 30R |
|  | 1a | 30T |
|  | 3a | 31F |
|  | 1a, 1b, 2a, 2b, 3a, 6a | 31M |
|  | 1a, 1b, 2a, 3a, 6a | 31V |
|  | 1a, 1b | 32del |
|  | 1a | 32L |
|  | 1a | 58D |
|  | 1a | 93C |
|  | 1a, 1b, 1c, 1h, 2a, 2b, 2d, 2j, 3, 3a, 4d | 93H |
|  | 1a, 1b, 4a | 93N |
|  | 1a | 93R |
|  | 1a | 93S |
|  | 1a | 93W |
| sofosbuvir | 1, 1a, 1b, 2, 3, 3a | 159F |
|  | 1a | 237G |
|  | 1a, 1b, 2a, 2b, 2d, 2j, 3, 3a, 4, 4a, 4r, 6a, 6l | 282T |
|  | 3 | 289I |
|  | 1b | 316H |
|  | 1b | 316N |
|  | 1a, 3, 3a | 321A |
|  | 1b, 4r | 321I |

#### **Supplementary Table 3: List of Resistance Associated Substitutions (RAS) considered relevant for each genotype.**

### Supplementary Figures

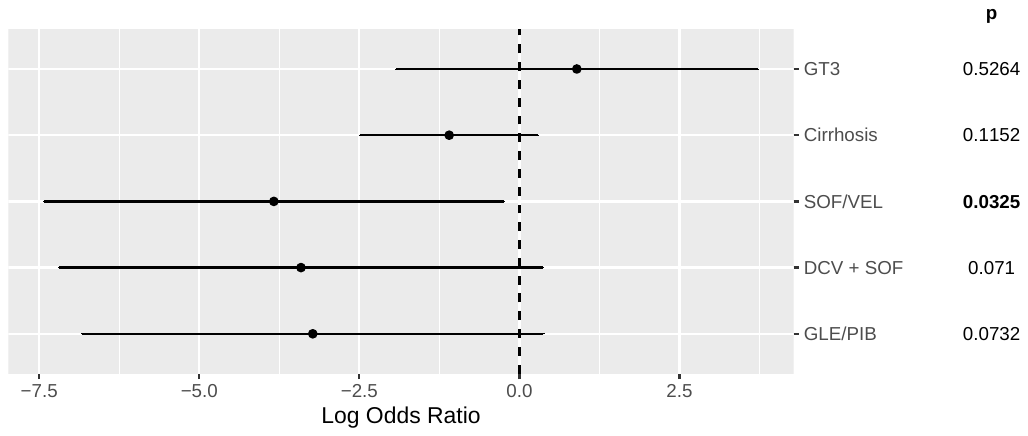

**Supplementary Figure 1: Multivariate analysis of clinical factors significantly associated with SVR or patients retreated with SOF/VEL/VOX according to baseline and treatment characteristics for the whole cohort (n=144).** The log odds ratio with 95% confidence intervals shown calculated using logistic regression. Values for which the SVR rate was 100% have not been included. Daclatasvir (DCV), Sofosbuvir (SOF), Glecaprevir (GLE), Pibrentasvir (PIB), Velpatasvir (VEL).

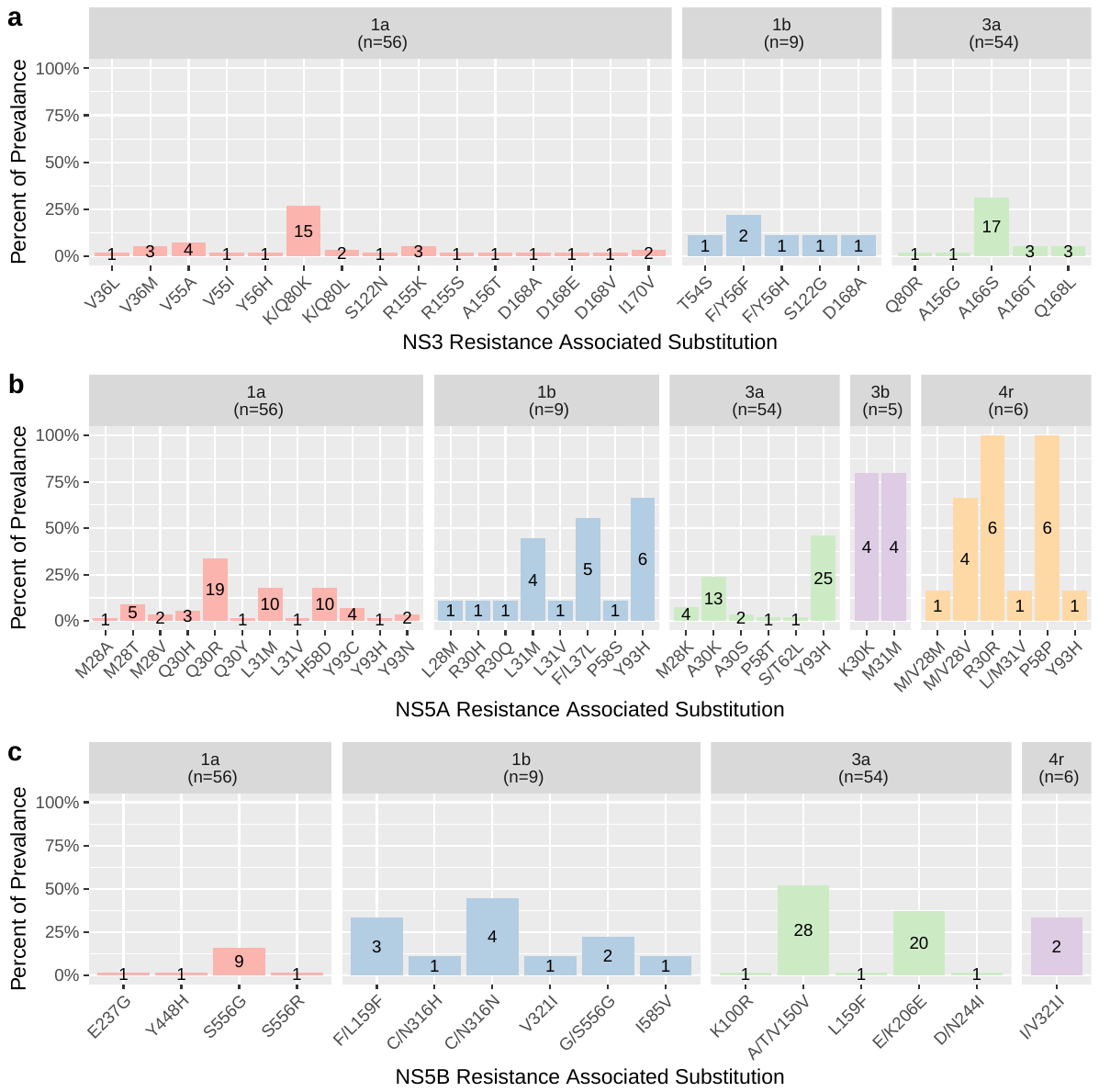

**Supplementary Figure 2: Prevalence of individual Resistance Associated Substitutions by HCV subtype. a) NS3 protein. b) NS5A protein. c) NS5B protein.** The percentage prevalence of each RAS detected is shown for each subtype with a frequency of more than two. The number of patients with the RAS shown in the bar.

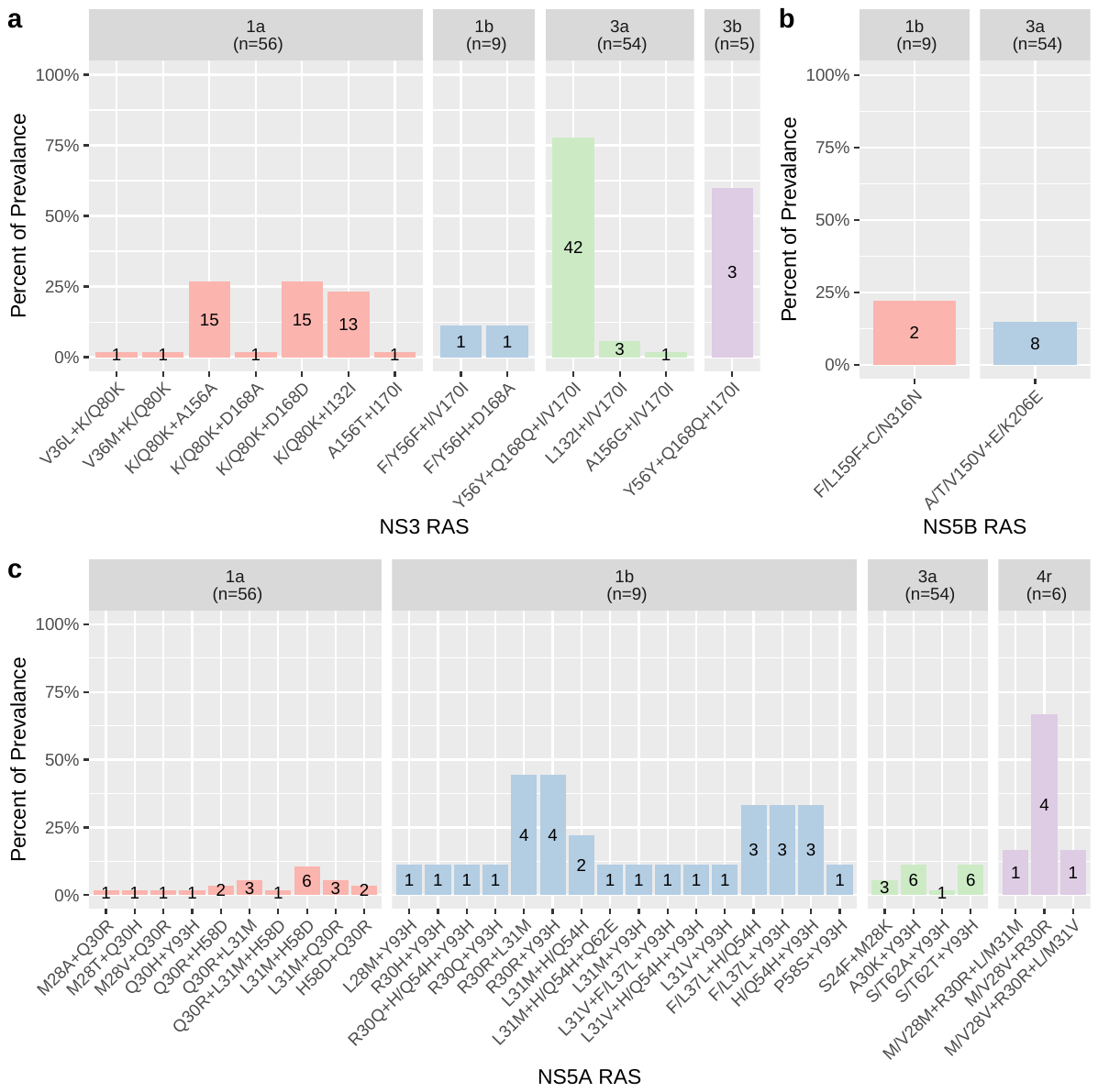

**Supplementary Figure 3: Prevalence of Resistance Associated Substitution combinations split by HCV subtype. a) NS3 Protein. b) NS5B protein. c) NS5A protein.** The percentage prevalence of each RAS detected is shown for each subtype with a frequency of more than two. The number of patients with the RAS shown in the bar.
